# DISCHARGE PLANNING PATIENT STROKE A NURSING SCIENCE PHILOSOPHY: SYSTEMATIC REVIEW

**DOI:** 10.1101/2023.12.07.23299664

**Authors:** Dwi Retnaningsih, Moses Glorino Rumambo Pandin, Nursalam Nursalam, Desi Ramadhani

## Abstract

**Objectives:** The quality of life and health of the patient are strongly influenced by the high incidence of stroke in a particular community or population. After the early recovery phase, recovery planning is crucial, as individuals who have suffered a stroke face uncertainty about the long-term recovery process as well as adjustments in everyday life. The objective of the research is to discuss the philosophical review of discharge planning for stroke patients from the points of view of ontology, epistemology, and axiology, with a focus on the philosophy of nursing science.

**Materials and methods:** In accordance with PRISMA (2017–2023) criteria, the study is a review of the literature. Keyword searches for “discharge planning,” “stroke,” “patients,” and “RCT” were conducted in the databases of Scopus, Science Direct, and ProQuest. The inclusion criteria encompassed quantitative research design, English-language publications, studies involving adult stroke patients, interventions related to discharge planning, and analysis evaluating discharge planning for stroke patients, yielding 281 identified papers. The Rayyan application is used in the selection process. PICO synthesis and grouping, which are based on the Cochrane Handbook for Systematic Review, are used in the analysis. Using the Joanna Briggs Institute Critical Assessment methodology, methodological quality assessment demonstrates that a multidisciplinary approach through ontological, epistemological, and axiological investigations offers a solid basis for enhancing health information systems and clinical practice.

**Results:** There are twenty-two quantitative research findings that show the features of the respondents, who were family caregivers and stroke patients. Ensuring that stroke victims receive the best care possible when they leave the hospital is the aim of recovery planning. In the process, families, patients, and health care providers collaborate. Patient assessment, treatment planning, medication scheduling, patient and family education, advanced care planning, mental and spiritual health support, health service coordination, medical recordkeeping, and monitoring are all essential elements of recovery planning.

**Conclusion:** Thorough recovery planning guarantees effective monitoring of medical records and coordination of treatment, improving the post-hospital experience for stroke patients. With an emphasis on the best possible recovery, this planning aims to offer focused and well-coordinated treatment. Stroke patients should anticipate comprehensive and well-coordinated post-hospital treatment, which will benefit both patients and families, thanks to the cooperation of health care professionals, patients, and families.

## INTRODUCTION

Stroke is one of the neurological diseases that has a significant impact on global public health. In addition to causing physical consequences, strokes can also result in cognitive and emotional changes that require special attention in post-stroke treatment planning. Deaths following a stroke have, over the previous 30 years, risen globally (Peng *et al*., 2023). Strokes can also occur because there is a relationship between environmental factors (Ranta *et al*., 2023). Discharge planning has become a critical element in efforts to coordinate the care of stroke patients after leaving the hospital environment. This phase not only covers the physical aspects but also considers the psychosocial and rehabilitation aspects of the patient. The findings of the study can provide information to researchers and policymakers who want to do discharge planning to improve hospitalization (Rachamin, Grischott and Neuner-Jehle, 2021).

The importance of nursing perspectives in discharge planning lies not only in the technical aspects of medical care but also in empowering families and patients during the healing process. Planning the recovery of stroke patients needs to be implemented and used by the health team in carrying out care and treatment (Simbolon *et al*., 2019; Sutin *et al*., 2022). A comprehensive method including several members of the health team, including nurses, doctors, physiotherapists, occupational therapists, and other health professionals, is recovery planning for stroke patients. Making sure the patient has a suitable and long-lasting treatment plan after leaving the hospital is the main goal of recovery planning.

The philosophy of nursing science provides a strong foundation for understanding the importance of holistic care and patient empowerment within the framework of post-stroke care. This literature review will cover key aspects of discharge planning, including patient needs assessment, patient and family education, short-term and long-term care planning, drug management, and psychosocial support. By integrating the philosophical perspective of nursing science, the research is expected to contribute to the development of a more holistic and patient-centred post-stroke care model (Smyth *et al*., 2023).

The study “Discharge Planning Patient Stroke: A Nursing Science Philosophy: Systematic Review,” which puts the philosophy of nursing science in perspective when analyzing stroke patient recovery planning systematically, adds innovation to the field since no prior research with similar issues has been done before. Understanding and raising the standard of care for stroke patients after they leave the hospital is made easier with this development. Through this approach, it is anticipated that study will yield new insights into the management of post-hospital stroke patient care, which in turn can improve patient health outcomes and overall quality of life. Although discharge planning is a major focus in post-stroke care, there is not much research that has comprehensively explored the philosophy of nursing science in this context. It is anticipated that the findings of this literature analysis will contribute to a better comprehension of the intricacy of discharge planning for stroke patients, as well as serve as a foundation for the creation of useful rules and regulations that enhance the standard of post-stroke treatment more successfully. In light of this, the study intends to conduct a thorough analysis of the literature on discharge planning for stroke patients, with a particular emphasis on nursing science’s charitable viewpoint.

## METHOD

### Inclusion and exclusion criteria

The inclusion and exclusion criteria are applied by the search and selection criteria. (1) Using PICOS (population, intervention, comparison, outcome, and study design) is one of the inclusion criteria. P stands for population, meaning research should involve adult-age stroke patients, Intervention is the use of all kinds of research in discharge planning in stroke patients. Discharge planning for stroke patients is an important process to ensure that patients receive adequate care after leaving the hospital and can reintegrate into the community. Discharge planning should involve collaboration between patients, families, and healthcare teams to ensure a smooth transition from hospital care to home or other facilities. This plan must be tailored to the unique needs of each stroke patient. C (comparison) is a comparative group for the discharge planning of stroke patients, O (outcome), which means the research includes analysis that supports the discharging planning of the stroke patient. All forms of quantitative study fall under research design (S);(2) the recent five years, from 2017 to 2023, are the year of publication; and (3) all of the publications are published in English. Protocol studies, conference presentations, editorial and review papers, case reports, case series, and applied design and development are among the criteria that are excluded.

### Search strategy

Research using literature review methods. The article review deals with the philosophical study of discharge planning stroke patients, examining axiological, bepistemological, and ontological viewpoints. The guidelines used for conducting systematic surveys are the Systematic Review and Meta-Analysis Preferred Reporting Items (PRISMA) (Page *et al*., 2021). The search for literature strategy is carried out by using keywords that match research topics such as “discharge planning,” “stroke,” “patients,” and “RCT” and searching articles through databases such as Scopus, Science Direct, and ProQuest. Journal articles were taken from 2019 to 2024. In this literature analysis research method, the main objective is to explore, analyze, and synthesize information from relevant literature to gain a deeper understanding of how discharge planning affects stroke patients, with a focus on the philosophical perspective of nursing science. The study was identified from the databases (N = 281), Science Direct (200), ProQuest (78), and Scopus. Using a combination of three major databases is the minimal requirement for a literature search in systematic surveys (Rethlefsen *et al*., 2021). The main search term is to use keywords that match the research topic: “discharge planning,” “stroke,” “patients,” “RCT,” combined with the boolean “AND/OR”.

### Selection of Studies

All authors (DR, DR, NN, and MG) scan the academic database. We next carried out the article selection procedure. Reviewers (DR, DR) choose publications based on Rayyan’s intelligent, systematic review. Rayyan is an app for mobile and online that facilitates systematic reviews (Ouzzani *et al*., 2016). Rayyan has proven to be effective in conducting systemic reviews and has significant potential for relieving the burden on the reviewer (Li *et al*., 2023). Three large databases’ worth of filtered articles were added to the Rayyan program. From this first database search, 281 searches were identified in all. After then, the articles were examined, and any duplicates were eliminated. After doing a thorough search, DR and DR separately go over the titles and abstracts found, and then they choose which papers meet the predetermined inclusion criteria. Titles and abstracts are filtered to include articles referring to cancer patients’ resilience. We found 22 relevant studies from major databases, read them independently in full text, and included them in this systematic review (Figure 1).

**Figure 1.1.**
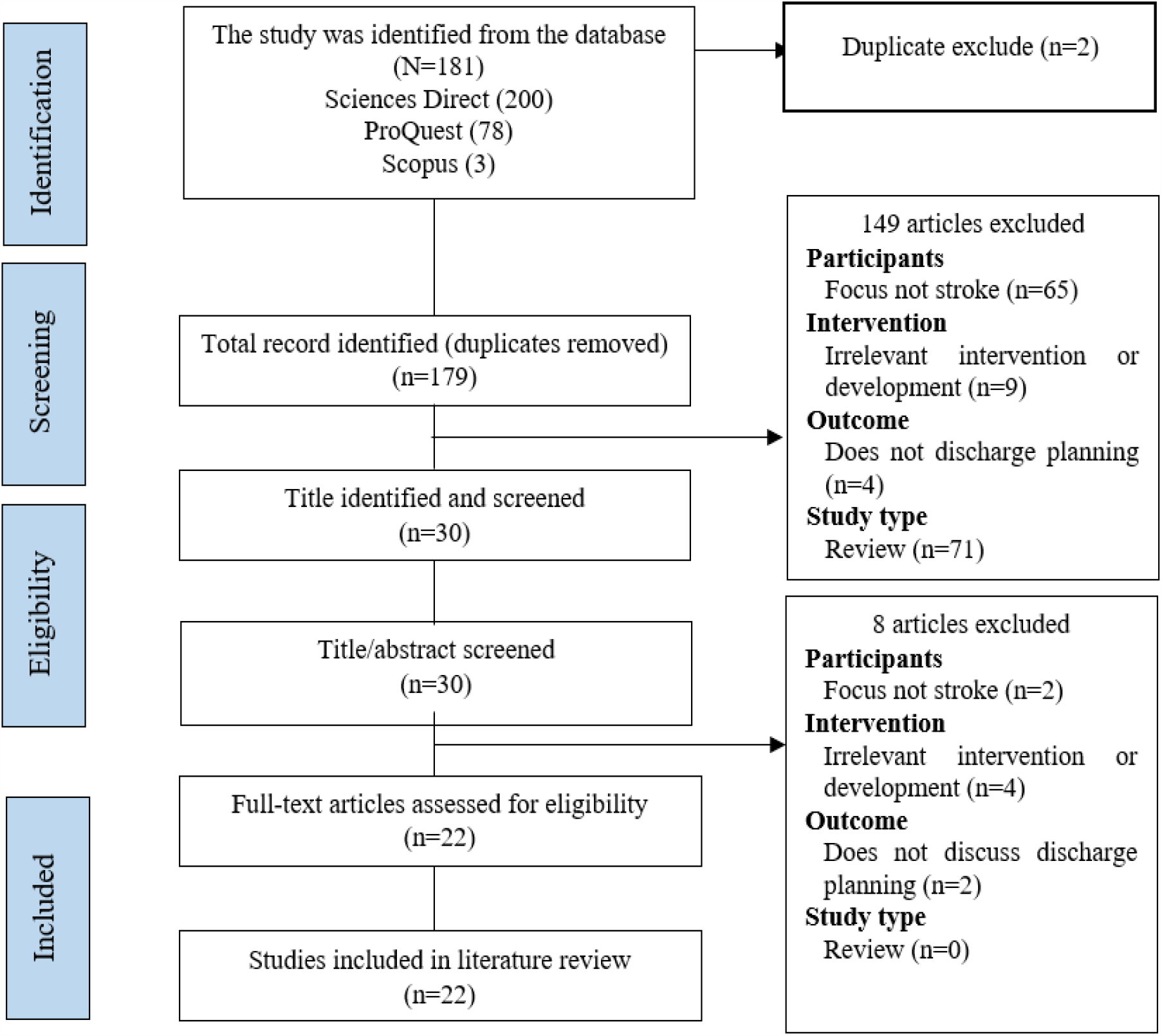
Process flow diagram for PRISMA literature search and filtering

### Risk of bias and research quality

PICO synthesis is used for analysis. Finding the study features (population, intervention, comparison, results, and research design) for each article acquired is the first step in the synthesis process. These are then grouped in accordance with the Cochrane Handbook for Systematic Review (Cumpston *et al*., 2021). The authors identify the quality of the research by considering the risk of bias. Assessment aims to assess the methodological quality of a study and determine to what extent a study has overcome the possibility of bias in its design, implementation, and analysis. To identify risk bias, systematic reviews use the Joanna Briggs Institute (JBI) critical assessment tool that follows the research design (Xie *et al*., 2023).

The researchers also used the search criteria of this review literature with the PRISMA flow diagram to obtain a suitable and eligible research article.

## RESULTS AND DISCUSSION

According to the results, a methodical approach to stroke patient recovery planning that incorporates nursing science philosophy offers a thorough understanding of the intricate medical and philosophical issues at play. This study provides a better and more thorough understanding of the needs of stroke patients after they leave the hospital by outlining holistic and sustainable recovery care plans. This strengthens the role of nursing science philosophy in this regard.

### 1. Ontological study of stroke patients’ discharge planning

The structure and hierarchy of concepts in the discharge planning process are analyzed in an ontological study of stroke victims. Ontology, as a branch of philosophy, examines the truth of existence and the relationship between entities. In the ontological study of discharge planning stroke patients, the focus is on an in-depth understanding of the basic concepts of exit planning after stroke. This research provides a conceptual basis for designing discharge planning for stroke patients from an ontological perspective. By understanding its ontological structure, health professionals can effectively integrate these concepts into clinical practice, improve the quality of stroke patient care, and support the development of more efficient health information systems.

Improving the quality of stroke patient care involves cross-sectoral cooperation, including medical personnel, patients, families, and health care facilities. This includes routine training for medical personnel, the formation of holistic care teams, the implementation of standard care protocols, routine evaluation and monitoring, integration of health technology, education for patients and families, coordinated rehabilitation programs (Gauthier *et al*., 2022), prevention of recurrences such as depression that may occur in stroke patients (Moriarty *et al*., 2020), patient participation in care planning, and building partnerships with community health care facilities as well as patient and family support (Jabal *et al*., 2024). As revealed in research by Phan and his colleagues (Phan *et al*., 2022), there is the first national picture of cross-sectional stroke services in Vietnam. Future research should involve systematic clinical audits of stroke treatments to validate data from the hospital. Repeat surveys in the coming years are expected to track progress and potentially affect increased stroke treatment capacity in Vietnam.

### 2. Epistemological study of stroke patients’ discharge planning

Epistemological research related to discharge planning in stroke patients will include an in-depth exploration of the nature of knowledge involved in the patient’s exit planning. Epistemology, as a branch of philosophy, explores the sources, limits, and foundations of knowledge. In the context of epistemological research on discharge planning in stroke patients, attention is focused on how knowledge related to the process is acquired, understood, and justified. Through an epistemological approach, we can gain insight into the origins and characteristics of knowledge in the context of discharge planning for stroke patients. It can enhance our understanding of how that knowledge is used and evaluated in health practice, as well as stimulate consideration of how to improve this process using epistemological approaches, through locating and researching the notion of stroke (Sato *et al*., 2020) and stroke patient care (Dimitriadis *et al*., 2022).

To acquire, understand, and validate knowledge related to the recovery planning process of a stroke patient, several steps can be taken: (1) Study the latest medical literature, books, journals, and research articles dealing with recovery plans for stroke patients in order to understand the latest scientific foundations and practices in stroke care. (2) Attend seminars, conferences, or workshops that focus on stroke management and recurrence planning to listen to experts, interact in person, and share experiences with other health professionals. (3) Consult with physicians, nurses, physiotherapists, occupational therapists, and other health professionals who have practical experience in treating stroke patients in order to gain practical insights and insights from their clinical experience. (4) Take advantage of reliable online resources such as health organization websites, medical institutions, or research centers that provide up-to-date and verified information on recovery planning for stroke patients. (5) Relates to guidelines and standards of care issued by national and international health institutions, like the World Health Organization (WHO) or the American Heart Association (AHA), which often offer guidelines for stroke treatment. (6) Take part in a training or certification program related to stroke management and recovery planning to gain a deeper understanding and updates related to best practices. (7) Join the health community, both online and offline, to discuss with other health professionals and exchange information to deepen understanding. (8) Follow study groups or online discussion forums that focus on stroke treatment and recovery planning topics to understand the various approaches and challenges faced by other health professionals. (9) Stay connected with the latest literature through subscriptions to health journals, newsletters, or professional social media. (10) Having acquired knowledge, apply these concepts in everyday practice, evaluate their effectiveness, and always be open to improvement and adaptation. It is important to combine information from different sources and always verify the reliability of the information received.

### 3. Axiological study of stroke patients’ discharge planning

The axiological study of discharge planning in stroke patients will involve examining the values and moral principles involved in the patient’s exit planning process. Axiology is a branch of philosophy that studies moral values. In the context of the study of the axiology of discharge planning for stroke patients, the focus will be on how these values influence and shape care practices. Identify the key values underlying discharge planning for stroke patients. It can include values such as patient autonomy, justice, humanity, trust, and certainty. analysis of how values influence interaction with patients and families during the planning process. how such values can affect communication, joint decision-making, and empower patients in planning their recovery. According to research carried out by Arab et al. (Arab *et al*., 2022), arguing that because of the involvement of patient privacy in home care situations, Professional identity perception, privacy, managing family interactions, shared security, and cultural-religious competency are just a few of the ethical values that become extremely important—even more important than hospital treatment.

The attitude of the health care staff and the family has an impact on the autonomy of the patient. Excessive protection, paternalism, routine care, and inconsistent approaches can hinder autonomy. On the contrary, attention, tailored intervention, and respectful dialogue can facilitate autonomy. This includes moderate instrumental and emotional support from the family government. Result-based multidisciplinary guidelines can raise attention to the autonomy of stroke patients and encourage team approaches (Proot *et al*., 2000).

The situation is different when the patient doesn’t have a home. Grech & Raeburn, in their research, identified five themes related to the provision of community-focused health services for the homeless. First off, the hospital serves as a safe haven for many homeless people. Second, nurses frequently believe that it is challenging to address the intricacy of the homeless health issue. Third, the stigma makes it difficult to provide community health care to homeless people living in hospitals. Fourth, granting the homeless the capacity to make decisions that are deemed significant. Fifth, there aren’t enough release choices or suitable connections between public services and hospitals. There are many obstacles in the way of providing hospital-based health services, and nurses’ perspectives shed light on what it’s like to care for the homeless on a daily basis in medical settings (Grech and Raeburn, 2021).

**Table 1.** The outcomes of the axiological perspective approach-based article review (n=22)

| No | Author | Method | Objective | Subject | Output |
| --- | --- | --- | --- | --- | --- |
| 1. | (Cadilhac *et al.*, 2017) | Mixed methods (explanatory | Create and evaluate multidisciplinary experiments, controlled pre- | Stroke patients and health workers | Improving patient return planning services in hospitals through a series of interventions involving several steps, |
|  |  | sequential design) | after-design observational studies, and organizational interventions to enhance recurrence therapy using stroke as a case study utilizing mixed techniques. |  | based on existing evidence, has proven effective and sustainable over a long period of time. Further testing of these interventions in other hospitals is thought to provide positive benefits. |
| 2. | (Duncan *et al.*, 2018) | Randomized control trial | The purpose of the study is to determine the effects of using COMPASS (Comprehensive Post-Acute Stroke Services) applications on post-stroke services and "Patient Reported Outcomes" (PRO), a tool for measuring stroke patients' health that involves direct patient reporting regarding their experiences with their post-stroke health. | Stroke patient | Two-thirds of clinicians who used COMPASS-CP expressed high levels of satisfaction with the tool, and three-quarters of doctors found significant factors affecting patients' recovery that may have gone unnoticed. |
| 3. | (Erler *et al.*, 2019) | Studi Kohort | Three months after leaving the rehabilitation nursing home for stroke patients, the independent contribution of perceived social support to involvement was examined. | Stroke patient | A unique factor contributing to the participation variations was the support received after being discharged from the hospital following an ischemic stroke. After three months. |
| 4. | (Westerlin d *et al.*, 2019) | Kuantitatif Studi Kohort Prospektif | Investigate the degree of return to work (RTW) post-stroke and whether very early cognitive function screening can predict return to work (RTW) after stroke. | Stroke patient | Both global cognitive function and executive function at 36–48 hours after stroke predict RTW levels at 6 or 18 months. Male sex, lower stroke severity, and not being on sick leave before a stroke are significant RTW tactics. |
| 5. | (Kaufman *et al.*, 2019) | Kuantitatif Studi Kohort Retrospektif | Evaluate the relationship of the Medicare Shared Savings Program (MSSP) with patient outcomes in the year after hospitalization for ischemic stroke. | Stroke patient | There's no connection between MSSP and recurring strokes. |
| 6. | (Kusuma Putri1, Rahayu and Agustina, 2019) | Cross-sectional | Identifying risk factors for stroke recurrence based on the Stroke Prognosis Instrument (SPI). | Stroke patient | There is a significant difference in risk factor scores between primary attack groups and recurrent strokes. Simple linear regression indicates that the incidence of stroke is positively correlated with the risk of recurring stroke. |
| 7. | (Muhsinin, Huriyah and Firmawati, 2019) | Quasi Experiment | Know the influence of the health education video project in the process of discharge planning to improve family readiness for treating stroke patients. | Stroke patient's family | Health education video projects in the process of discharge planning influence improving family readiness when treating stroke patients at home. |
| 8. | (Jones, McClean and Stanford, 2019) | Kuantitatif | Identifying old models of hospitalization after stroke and how to prepare for recovery using the stroke recovery phase model | Stroke patient | This model has obvious practical value as it produces a long distribution of nursing care based on age and type of stroke, which is useful in home planning. |
| 9. | (Mohammadi *et al.*, 2019) | Randomized control trial | Creating discharge planning programs for Iranian stroke patients' relatives who | Stroke patient's family | In the group trial, there was a statistically significant increase in parental readiness. Stress levels were |
|  |  |  | provide care. as well as assessing the program's effectiveness in relation to their stress levels and level of foster care readiness. |  | lower in the experimental group than in the control group. |
| 10 | (Sheehy *et al.*, 2020) | Randomised controlled trial | presents the challenges and solutions that arise during RCT, one intermediate location of rehabilitation interventions carried out in the hospital stroke rehabilitation series. | Stroke patient | We have identified the challenges in providing rehabilitation interventions to stroke patients and some of the actions taken to address these challenges. Through good planning, flexibility, and collaboration, almost all of the challenges have been overcome. |
| 11 | (Tan *et al.*, 2020) | Randomized control trial | Examine how continued nursing care affects ischemic stroke patients' chances of recovery. | Stroke patient | Treatment compliance, quality of life ratings, self-rating depression scales (SDS), self-rating anxiety scales (SAS), and the National Institutes of Health Stroke Scale differ between the control and intervention groups (NIHSS). |
| 12 | (Tanlaka *et al.*, 2020) | Studi Kohort | Evaluate gender differences in stroke rehabilitation results. | Stroke patient | Age and gender have a major impact on the length of stay (LOS) that stroke patients require for rehabilitation and recovery. These variations would suggest that treatment planning should take the patient's age and gender into consideration. |
| 13 | (Said Taha and Ali Ibrahim, 2020) | Quasy experiment | Analyze how recovery planning strategies for stroke patients affect daily activities and quality of life. | Stroke patient | Patient knowledge, quality of life, and daily activities are positively correlated, with statistically significant differences between them. |
| 14 | (Ottiger *et al.*, 2020) | Studi Kohort | determining the ADL boundary scores that enable doctors to decide whether stroke patients who are "living alone" or "living with family" can be returned home. Or you have to go to the nursery. | Stroke patient | stroke-crossers who live alone require a higher rate of ADL to return home than those who live with families. |
| 15 | (Farahani *et al.*, 2020) | Kohort longitudinal | Knowing the nursing needs of the elderly stroke patient's family. | The family of a stroke patient | Two weeks after recovery and twelve weeks after return, there is a strong association between the number of family nursing needs and the age at which the old patient survives. Pre-discharge physiotherapy patients and their caregivers are related, as is the duration of the patient's hospital stay and the number of caregivers required three months after the patient returns. |
| 16 | (De Silva, Tan and Thilarajah, 2020) | Kuantitatif | To use and ensure that key elements of an acute stroke unit are | Stroke patient | Three categories exist: stroke complications (concentrating on five common complications), unified teams (describes daily check-in, patient education, communication, return planning, and post-refoulement support), and acute monitoring (neurological observation, blood, pressure monitoring, input-output, investigation, and post-reperfusion-specific problems). |
| 17 | (Lin *et al.*, 2021) | Randomised controlled trial | Examine the effects of a health education program taught by nurses on stroke patients and family nurses during the transition from hospital to home care. | a stroke patient and a babysitter. | Programs for nurse-led health education enhance the health of stroke survivors and their nurses. |
| 18 | (Mandigout *et al.*, 2021) | Randomized control trial | Assess the impact of the six-month individualized home-building program on the ability to walk, as measured by a six-minute walking test, in patients with subacute stroke. | Stroke patient | Following a customized training program that included home visits, there was no difference in the six-minute walking test between the two groups of subacute stroke patients six months later. |
| 19 | (Xu, Liu and Zhang, 2021) | Kuantitatif experiment | Gathering relevant data and investigating therapeutic approaches for level 3 rehabilitation in particular Chinese locales. | Stroke patient | There was an increase in Fugl-Meyer Analysis (FMA) and Modified Barthel Index MBI scores from both groups after treatment. |
| 20 | (de Berker *et al.*, 2021) | Kuantitatif | Describe the relationship between pre-stroke defects, stroke severity, and the goal of recovery to develop a predictive model of the goal of recovery that can be understood and trusted by doctors. | Stroke patient | The Modified Rankin Scale (mRS) is the strongest predictor for the purposes of patient return. The National Institutes of Health Stroke Scale (NIHSS) is only predictive when combined with other variables. |
| 21 | (Aoki and Nakayama, 2022) | Randomized control trial | Evaluate the impact of the use of decision-making aids (DAs) on participation in decision-making and conflicts regarding post-return care selection and location for stroke victims who are older adults and their families. | Stroke patient | The decision conflict scale (DCS) and the control preference scale revision (CPS), respectively, will be used to assess the primary (decision-making conflict) and secondary (decision-making participation) findings. A double-directional repeat measurement z test and variance analysis will be used, respectively, to examine intergroup differences in the DCS and CPS. |
| 22 | (Gauthier *et al.*, 2022) | Randomized control trial | Integrating behavioral interventions into motor rehabilitation | Stroke patient | Interpretation The outcomes of self-managed motor games combined with telehealth behavioral sessions are comparable to those of CI in therapy clinics. |

### 4. Stroke patients’ discharge planning

Making plans for stroke victims’ discharge is a structured process to ensure that patients get proper and supportive treatment after they leave the hospital. It involves collaboration between health care teams, patients, and families to ensure a smooth transition from inpatient care to home or other environmental care. Here are some common components of discharge planning for stroke patients: (1) A study of the patient’s medical history and the determination of whether long-term care or rehabilitation is required are included in the patient condition assessment (Geerars, Wondergem and Pisters, 2021). (2) Rehabilitation planning by identifying the type of rehabilitation therapy required, such as physiotherapy, occupational therapy, and logopedics, appointments, and rehabilitation programs at the rehabilitation center or at home. (3) Medicines to ensure a clear understanding of dosage, frequency, and side effects (Firth *et al*., 2023). (4) Educate patients and families by providing information on stroke conditions, warning symptoms, actions to be taken, instructions on lifestyle changes, diet, and recommended exercise (Williamson *et al*., 2021; Sim and Shin, 2024). (5) Mental health support by assessing the mental health support needs of patients and families (Turi *et al*., 2023) as well as spiritual support (Yousofvand *et al*., 2023) and reference to mental health services or psychosocial support if necessary. (6) Advanced care is determined by determining whether a patient needs advanced care or follow-up by a particular health professional. Arrange appointments and follow-ups with a doctor or other health professional (Lin *et al*., 2020). (7) Coordination of health services with other health service providers involved in patient care. Ensure that relevant medical information is shared with all relevant service providers (Eastman, Kalesnikava and Mezuk, 2022). (8) Monitoring documentation and information by ensuring complete and accurate medical records. Provide information on how to monitor symptoms or warning signs requiring immediate medical attention (Fernandes *et al*., 2021).

In order to guarantee continued treatment and the best possible recovery following strokes, health care professionals, patients, and families must work together during the discharge planning process (Lin *et al*., 2022; Hyvärinen *et al*., 2023).

## CONCLUSION

This study concludes that a solid foundation for enhancing clinical practice and health information systems is provided by comprehensive recovery planning, which incorporates philosophical methods of ontology, epistemology, and axiology from the standpoint of nursing science. This study, which focuses on the best possible outcome, demonstrates that organizing the recuperation of stroke victims entails not only evaluating their physical state but also considering elements of rehabilitation, providing support for their mental and spiritual well-being, and organizing medical resources. The findings demonstrate how important it is for patients, families, and healthcare providers to work together to provide coordinated and thorough post-hospital care. This research suggests that stroke patients and their families may benefit from a more positive post-hospital experience through the use of holistic recovery planning.

## Data Availability

All data produced in the present work are contained in the manuscript

## ACKNOWLEDGEMENT

For providing the facilities for this study, the authors are grateful to the Faculty of Nursing at Universitas Airlangga.

## REFERENCE

Aoki, Y. and Nakayama, K. (2022) ‘Improving older adults stroke survivors decisionlmaking when selecting a discharge.pdf’, International Journal of Nursing Knowledge, 34, pp. 185–192. Available at: DOI: 10.1111/2047-3095.12393.

Arab, M. et al.x (2022) ‘Nurses’ experiences of the ethical values of home care nursing: A qualitative study’, International Journal of Nursing Sciences, 9(3), pp. 364–372. Available at: 10.1016/j.ijnss.2022.06.008.

de Berker, H. et al.x (2021) ‘Pre-stroke disability and stroke severity as predictors of discharge destination from an acute stroke ward’, Clinical Medicine, Journal of the Royal College of Physicians of London, 21(2), pp. E186–E191. Available at: 10.7861/CLINMED.2020-0834.

Cadilhac, D.A. et al.x (2017) ‘Improving discharge care: The potential of a new organisational intervention to improve discharge after hospitalisation for acute stroke, a controlled before-after pilot study’, BMJ Open, 7(8), pp. 1–10. Available at: 10.1136/bmjopen-2017-016010.

Cumpston, M.S. et al.x (2021) ‘The use of “PICO for synthesis” and methods for synthesis without meta-analysis: protocol for a survey of current practice in systematic reviews of health interventions’, F1000Research, 9, pp. 1–9. Available at: 10.12688/F1000RESEARCH.24469.1.

Dimitriadis, K. et al.x (2022) ‘Moving from traditional to more advanced treatments in stroke care is cost-effective: A case study from Greece’, Journal of Stroke and Cerebrovascular Diseases, 31(11), p. 106764. Available at: 10.1016/j.jstrokecerebrovasdis.2022.106764.

Duncan, P.W. et al.x (2018) ‘COMPASS-CP: An Electronic Application to Capture Patient-Reported Outcomes to Develop Actionable Stroke and Transient Ischemic Attack Care Plans’, Circulation. Cardiovascular quality and outcomes, 11(8), pp. 1–11. Available at: 10.1161/CIRCOUTCOMES.117.004444.

Eastman, M.R., Kalesnikava, V.A. and Mezuk, B. (2022) ‘Experiences of care coordination among older adults in the United States: Evidence from the Health and Retirement Study’, Patient Education and Counseling, 105(7), pp. 2429–2435. Available at: 10.1016/j.pec.2022.03.015.

Erler, K.S. et al.x (2019) ‘Social Support as a predictor of community participation after stroke’, Frontiers in Neurology, 10(September), pp. 1–7. Available at: 10.3389/fneur.2019.01013.

Farahani, M.A. et al.x (2020) ‘Investigating the needs of family caregivers of older stroke patients: A longitudinal study in Iran’, BMC Geriatrics, 20(1), pp. 1–12. Available at: 10.1186/s12877-020-01670-0.

Fernandes, L. et al.x (2021) ‘How to Improve Emergency Information Systems to Optimize the Care of Acute Stroke’, Procedia Computer Science, 196(2021), pp. 606–614. Available at: 10.1016/j.procs.2021.12.055.

Firth, N. et al.x (2023) ‘Stroke survivors’ perspectives on decision-making about rehabilitation and the prospect of taking recovery-promoting drugs: A qualitative study’, Exploratory Research in Clinical and Social Pharmacy, 11(April), p. 100297. Available at: 10.1016/j.rcsop.2023.100297.

Gauthier, L. V. et al.x (2022) ‘Video game rehabilitation for outpatient stroke (VIGoROUS): A multi-site randomized controlled trial of in-home, self-managed, upper-extremity therapy’, eClinicalMedicine, 43, pp. 1–12. Available at: 10.1016/j.eclinm.2021.101239.

Geerars, M., Wondergem, R. and Pisters, M.F. (2021) ‘Decision-Making on Referral to Primary Care Physiotherapy After Inpatient Stroke Rehabilitation’, Journal of Stroke and Cerebrovascular Diseases, 30(5), p. 105667. Available at: 10.1016/j.jstrokecerebrovasdis.2021.105667.

Grech, E. and Raeburn, T. (2021) ‘Perceptions of hospital-based Registered Nurses of care and discharge planning for people who are homeless: A qualitative study’, Collegian, 28(1), pp. 1–9. Available at: 10.1016/j.colegn.2020.02.004.

Hyvärinen, S. et al.x (2023) ‘Healthcare professionals ‘experience regarding competencies in specialized and primary stroke unitsl: A qualitative study’, (xxxx). Available at: 10.1016/j.jvn.2023.11.006.

Jabal, M.S. et al.x (2024) ‘Machine learning prediction of hospital discharge disposition for inpatients with acute ischemic stroke following mechanical thrombectomy in the United States’, Journal of Stroke and Cerebrovascular Diseases, 33(1), p. 107489. Available at: 10.1016/j.jstrokecerebrovasdis.2023.107489.

Jones, B., McClean, S. and Stanford, D. (2019) ‘Modelling mortality and discharge of hospitalized stroke patients using a phase-type recovery model’, Health Care Management Science, 22(4), pp. 570–588. Available at: 10.1007/s10729-018-9446-6.

Kaufman, B.G. et al.x (2019) ‘The Medicare Shared Savings Program and Outcomes for Ischemic Stroke Patients: a Retrospective Cohort Study’, Journal of General Internal Medicine, 34(12), pp. 2740–2748. Available at: 10.1007/s11606-019-05283-1.

Kusuma Putri1, T.A.R., Rahayu, L.P. and Agustina, E.N. (2019) ‘Stroke Recurrence Based on Stroke Prognosis Instrument II (SPI-II) and The Attack Number of Stroke’, KnE Life Sciences, 2019, pp. 923–930. Available at: 10.18502/kls.v4i13.5352.

Li, J. et al.x (2023) ‘Sensitivity and specificity of alternative screening methods for systematic reviews using text mining tools’, Journal of Clinical Epidemiology, 162, pp. 72–80. Available at: 10.1016/j.jclinepi.2023.07.010.

Lin, S. et al.x (2020) ‘The effect of transition care interventions incorporating health coaching strategies for stroke survivors: A systematic review and meta-analysis’, Patient Education and Counseling, 103(10), pp. 2039–2060. Available at: 10.1016/j.pec.2020.05.006.

Lin, S. et al.x (2021) ‘Nurse-led health coaching programme to improve hospital-to-home transitional care for stroke survivors: A randomised controlled trial’, Patient Education and Counseling, 105(4), pp. 917–925. Available at: 10.1016/j.pec.2021.07.020.

Lin, S. et al.x (2022) ‘The experience of stroke survivors and caregivers during hospital-to-home transitional care: A qualitative longitudinal study’, International Journal of Nursing Studies, 130, p. 104213. Available at: 10.1016/j.ijnurstu.2022.104213.

Mandigout, S. et al.x (2021) ‘Effect of individualized coaching at home on walking capacity in subacute stroke patients: A randomized controlled trial (Ticaa’dom)’, Annals of Physical and Rehabilitation Medicine, 64(4), p. 101453. Available at: 10.1016/j.rehab.2020.11.001.

Mohammadi, S. et al.x (2019) ‘The effect of family-oriented discharge program on the level of preparedness for care-giving and stress experienced by the family of stroke survivors’, Iranian Rehabilitation Journal, 17(2), pp. 113–120. Available at: 10.32598/irj.17.2.113.

Moriarty, A.S. et al.x (2020) ‘The role of relapse prevention for depression in collaborative care: A systematic review’, Journal of Affective Disorders, 265(November 2019), pp. 618–644. Available at: 10.1016/j.jad.2019.11.105.

Muhsinin, S.Z., Huriah, T. and Firmawati, E. (2019) ‘Health eduucation video project in discharge planning process to improves family preparedness in caring for stroke patients’, JHeS (Journal of Health Studies), 3(1), pp. 80–87. Available at: 10.31101/jhes.492.

Ottiger, B. et al.x (2020) ‘Can I Discharge My Stroke Patient Home After Inpatient Neurorehabilitation? LIMOS Cut-Off Scores for Stroke Patients “Living Alone” and “Living With Family”‘, Frontiers in Neurology, 11(November), pp. 1–10. Available at: 10.3389/fneur.2020.601725.

Ouzzani, M. et al.x (2016) ‘Rayyan-a web and mobile app for systematic reviews’, Systematic Reviews, 5(1), pp. 1–10. Available at: 10.1186/s13643-016-0384-4.

Page, M.J. et al.x (2021) ‘The PRISMA 2020 statement: An updated guideline for reporting systematic reviews’, The BMJ, 372. Available at: 10.1136/bmj.n71.

Peng, S. et al.x (2023) ‘Global, regional, and national time trends in mortality for stroke, 1990– 2019: An age-period-cohort analysis for the global burden of disease 2019 study and implications for stroke prevention’, International Journal of Cardiology, 383(April), pp. 117–131. Available at: 10.1016/j.ijcard.2023.05.001.

Phan, H.T. et al.x (2022) ‘Organisational survey for acute stroke care in Vietnam: Regional Collaboration Programme’, Journal of Stroke and Cerebrovascular Diseases, 31(11), pp. 1–7. Available at: 10.1016/j.jstrokecerebrovasdis.2022.106792.

Proot, I.M. et al.x (2000) ‘Stroke patients’ needs and experiences regarding autonomy at discharge from nursing home’, Patient Education and Counseling, 41(3), pp. 275–283. Available at: 10.1016/S0738-3991(99)00113-5.

Rachamin, Y., Grischott, T. and Neuner-Jehle, S. (2021) ‘Implementation of a complex intervention to improve hospital discharge: process evaluation of a cluster randomised controlled trial’, BMJ Open, 11(5), pp. 1–12. Available at: 10.1136/bmjopen-2021-049872.

Ranta, A. et al.x (2023) ‘Environmental factors and stroke: Risk and prevention’, Journal of the Neurological Sciences, 454(March), p. 120860. Available at: 10.1016/j.jns.2023.120860.

Rethlefsen, M.L. et al.x (2021) ‘PRISMA-S: an extension to the PRISMA Statement for Reporting Literature Searches in Systematic Reviews’, Systematic Reviews, 10(1), pp. 1–19. Available at: 10.1186/s13643-020-01542-z.

Said Taha, A. and Ali Ibrahim, R. (2020) ‘Effect of a Design Discharge Planning Program for Stroke Patients on Their Quality of Life and Activity of Daily Living’, International Journal of Studies in Nursing, 5(1), pp. 64–86. Available at: 10.20849/ijsn.v5i1.724.

Sato, D.M.V. et al.x (2020) ‘Ischemic stroke: Process perspective, clinical and profile characteristics, and external factors’, Journal of Biomedical Informatics, 111(September), p. 103582. Available at: 10.1016/j.jbi.2020.103582.

Sheehy, L. et al.x (2020) ‘Implementation of a randomized controlled trial on an inpatient stroke rehabilitation unit: Lessons learned’, Contemporary Clinical Trials Communications, 18, p. 100563. Available at: 10.1016/j.conctc.2020.100563.

De Silva, D.A., Tan, I.F. and Thilarajah, S. (2020) ‘A protocol for acute stroke unit care during the COVID-19 pandemic’, Journal of Stroke and Cerebrovascular Diseases, 29(9), p. 105009. Available at: 10.1016/j.jstrokecerebrovasdis.2020.105009.

Sim, J. and Shin, C. (2024) ‘Two stroke education programs designed for older adults’, Geriatric Nursing, 55, pp. 105–111. Available at: 10.1016/j.gerinurse.2023.10.014.

Simbolon, S. et al.x (2019) ‘The effectiveness of discharge planning stroke patient due to hypertension to improve patient satisfaction and independence’, Enfermeria Clinica, 29(Insc 2018), pp. 703–708. Available at: 10.1016/j.enfcli.2019.06.011.

Smyth, C. et al.x (2023) ‘To assess the effects of cross-education on strength and motor function in post stroke rehabilitation: a systematic literature review and metaanalysis’, Physiotherapy (United Kingdom), 119, pp. 80–88. Available at: 10.1016/j.physio.2023.02.001.

Sutin, U. et al.x (2022) ‘Problems and needs when caring for stroke patient at homes’, International Journal of Public Health Science, 11(2), pp. 695–705. Available at: 10.11591/ijphs.v11i2.21013.

Tan, S.-Y. et al.x (2020) ‘Application Study of Continuous Nursing Intervention among Ischemic Stroke Patients’, International Journal of Clinical and Experimental Medicine Research, 4(3), pp. 107–111. Available at: 10.26855/ijcemr.2020.07.013.

Tanlaka, E. et al.x (2020) ‘Sex Differences in Stroke Rehabilitation Care in Alberta’, Canadian Journal of Neurological Sciences, 47(4), pp. 494–503. Available at: 10.1017/cjn.2020.53.

Turi, E. et al.x (2023) ‘The effectiveness of nurse practitioner care for patients with mental health conditions in primary care settings: A systematic review’, Nursing Outlook, 71(4), p. 101995. Available at: 10.1016/j.outlook.2023.101995.

Westerlind, E. et al.x (2019) ‘Very early cognitive screening and return to work after stroke’, Topics in Stroke Rehabilitation, 26(8), pp. 602–607. Available at: 10.1080/10749357.2019.1645440.

Williamson, T.M. et al.x (2021) ‘Promoting adherence to physical activity among individuals with cardiovascular disease using behavioral counseling: A theory and research-based primer for health care professionals’, Progress in Cardiovascular Diseases, 64, pp. 41–54. Available at: 10.1016/j.pcad.2020.12.007.

Xie, C. et al.x (2023) ‘Long-term trend of future Cancer onset: A model-based prediction of Cancer incidence and onset age by region and gender.’, Preventive Medicine, 177(October), p. 107775. Available at: 10.1016/j.ypmed.2023.107775.

Xu, W., Liu, L. and Zhang, J. (2021) ‘Application Analysis Based on Big Data Technology in Stroke Rehabilitation Nursing’, Journal of Healthcare Engineering, 2021, pp. 1–10. Available at: 10.1155/2021/3081549.

Yousofvand, V. et al.x (2023) ‘Impact of a spiritual care program on the sleep quality and spiritual health of Muslim stroke patients: A randomized controlled trial’, Complementary Therapies in Medicine, 77(August), p. 102981. Available at: 10.1016/j.ctim.2023.102981.

